# Reprocessing N95 Respirators During the COVID-19 Pandemic: Moist Heat Inactivates SARS-CoV-2 and Maintains N95 Filtration

**DOI:** 10.1101/2020.05.25.20112615

**Authors:** Simeon C. Daeschler, Niclas Manson, Kariym Joachim, Alex W. H. Chin, Katelyn Chan, Paul Z. Chen, Kiana Tajdaran, Kaveh Mirmoeini, Jennifer J. Zhang, Jason T. Maynes, Michelle Science, Ali Darbandi, Derek Stephens, Leo L. M. Poon, Frank Gu, Gregory H. Borschel

## Abstract

**Background:** The unprecedented demand and consequent global shortage of N95 respirators during the COVID-19 pandemic have left frontline workers vulnerable to infection. To potentially expand the supply, we validated a rapidly applicable low-cost decontamination protocol in compliance with regulatory standards to enable the safe reuse of personalized, disposable N95-respirators.

**Methods:** Four common models of N95-respirators were disinfected for 60 minutes at 70°C either at 0% or 50% relative humidity (RH). Effective inactivation of SARS-CoV-2 and *E. coli* was evaluated in inoculated masks. The N95 filter integrity was examined with scanning electron microscopy. The protective function of disinfected N95 respirators was tested against US NIOSH standards for particle filtration efficiency, breathing resistance and respirator fit.

**Results:** A single heat treatment inactivated both SARS-CoV-2 (undetectable, detection limit: 100 TCID50/ml) and *E. coli* (0 colonies at 50%RH) in all four respirator models. Even N95-respirators that underwent ten decontamination cycles maintained their integrity and met US-governmental criteria for approval regarding fit, filtration efficiency and breathing resistance. Scanning electron microscopy demonstrated maintained N95 fiber diameter compared to baseline.

**Interpretation:** Thermal disinfection enables large-scale, low cost decontamination of existing N95 respirators using commonly sourced equipment during the COVID-19 pandemic. This process could be used in hospitals and long term care facilities and also provides a feasible approach to expand the N95 supply in low- and middle-income regions.

## Introduction

As the COVID-19 pandemic has overwhelmed many health care systems world-wide, the unprecedented demand for personal protective equipment exhausted stockpiles and interrupted global supply chains for N95 respirators. Currently, the proportion of frontline healthcare workers among SARS-CoV-2 infected individuals exceeds 10% in some regions and is expected to increase if stockpiles further diminish (1, 2). As a result, protecting frontline workers from SARS-CoV-2 infection is now an immediate global concern (3).

Disposable N95 respirators protect users against infectious airborne particles and are therefore critical to frontline workers during the COVID-19 pandemic (3). However, the present global shortage of personal protective equipment has forced regulating institutions to adjust infection control measures: prior to the pandemic, guidelines recommended disposal of N95 respirators after each patient encounter. Now, evolving guidelines instruct staff to re-use one mask over their whole shift or even longer (4). This policy of re-using disposable masks in areas of high airborne pathogen exposure, such as aerosol generating medical procedures in COVID-19 patient care, may result in accumulation of contagious material on the mask surface, risking the health and safety of personnel and patients (5, 6). Inactivating accumulated pathogens in disposable respirators without affecting their protective properties may enable safe reuse and thus help to alleviate the current global shortage temporarily. However, the sterilization methods regularly used in health care institutions potentially degrade disposable respirators and thereby affect fit or filtration efficiency (7).

Thermal disinfection may overcome this issue and potentially provides a widely available and cost-effective decontamination strategy for disposable respirators. Recent reports demonstrate a high sensitivity to heat for SARS-CoV-2 as five minutes of heating at 70°C inactivates the virus (5, 8). The polypropylene microfibers in commercially available N95 respirators have a thermal degradation point above 130°C, suggesting that the filter may withstand repetitive exposure to 70°C (9, 10). However, the viricidal efficacy of thermal disinfection for SARS-CoV-2-contaminated N95 respirators, and the protective performance of heat-treated respirators has not been validated to a level meeting Unites States (US) regulatory standards.

We therefore investigated whether thermal disinfection at 70°C for 60 min inactivates pathogens, including SARS-CoV-2, while maintaining critical N95 respirator protective properties for multiple cycles of disinfection and re-use in a real-world setting.

## Methods

### Thermal disinfection protocol for N95 respirators

We used thermal disinfection in cycles of 60 min at 70°C, either at 0% or 50% relative humidity (RH), to treat four common models of commercially available N95 respirators (8110s, 9105s, 8210, 1860s; 3M, USA). The respirators were wrapped in sterilization pouches (Steril-peel, GS Medical Packaging, Canada) prior to disinfection. To control for temperature and RH, we set the BevLes Heated Holding Cabinet with humidity (BevLes Inc, Erie, USA) to 70°C and varied humidity between 0% to 50% RH. A digital thermo- and hygrometer (Hadgen Group Inc, Canada) was used as an added quality control measure. Additionally, we accounted for potential real-world temperature fluctuations by cooling the masks to room temperature for five minutes mid-cycle.

### SARS-CoV-2 inactivation

We assessed SARS-CoV-2 inactivation in all four N95 respirator models. We cut unprocessed and 10x heat-treated N95 respirators into 1 cm^2^ pieces and inoculated the outer surface of the respirators with 5 µl of SARS-CoV-2 (∼7.8 log Fifty-percent tissue culture infective dose per ml [TCID50/ml] in triplicates (n=3 per respirator type) in a biosafety level 3 laboratory. The virus-inoculated respirators underwent thermal disinfection at 70°C at 0% RH for 60 min, with and without a 5 min cool down midcycle, followed by soaking in 300 µl of viral transport medium for 30 min for virus elution. We then titrated the recovered infectious virus particles by standard TCID50 assay using Vero E6 cells as described (5). Virus-inoculated respirator surfaces without the heat inactivation step were used as controls.

### Bacterial inactivation

To test for bacterial inactivation, we cut unprocessed N95 respirators (1860S, 3M, Minnesota, USA) into 1 cm^2^ pieces. The outer surface was inoculated with 100 µl of *Escherichia coli* (4 × 10^8^ CFU/ml, optical density 0.612 at 600 nm) and a negative control was inoculated with pure Luria-Bertani (LB) medium. The inoculated respirators underwent 60 min heat treatment at 70°C either at 0% RH or at 25% RH, 40% RH or 50% RH (n=4 per condition). Positive controls were treated at 90°C / 70% RH and negative controls were left at room temperature for one hour. We then washed N95 fragments individually in 1 ml of LB medium and inoculated 100 µl washing media on LB-agar plates. Colonies were counted after 24 h incubation at 37°C (Figure 1). To the 900µl of remaining washing media and N95 fragments, 9.1ml LB media were added and incubated at 37°C in a shaking incubator. Optical density at 600nm was read after 24 hours of incubation to estimate bacteria concentration.

**Figure 1:**
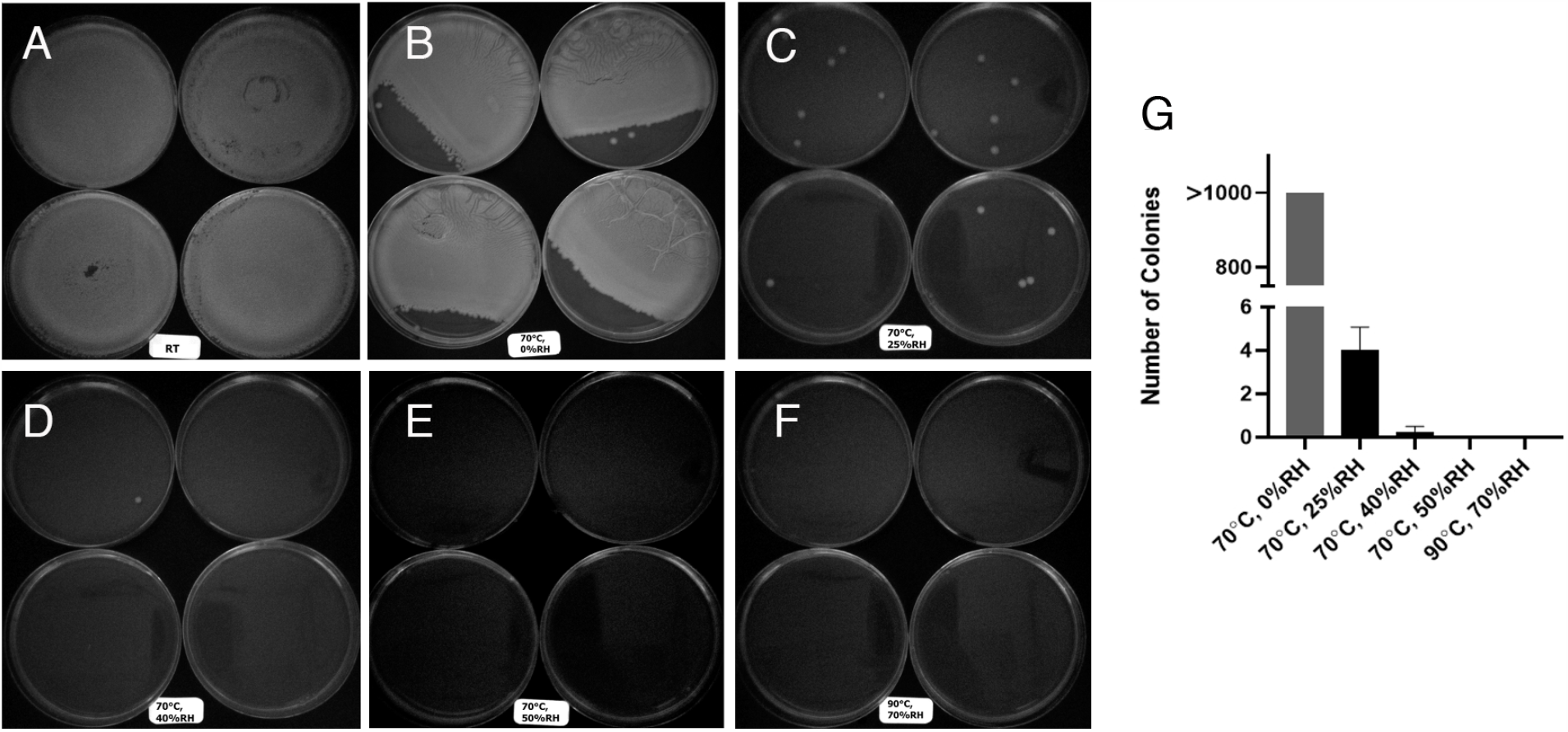
Bacterial inactivation in thermally disinfected N95 respirators. Panels A – F: *E. coli* colonies after 24h incubation at 37°C derived from inoculated N95 respirators that underwent treatment at different temperatures and humidity (A: incubation at room temperature (RT), B: 70°C / 0% relative humidity [RH], C: 70°C / 25% RH, D: 70°C / 40% RH, E: 70°C / 50% RH and F: 90°C / 70% RH as a control). Panel G: The *E. coli* colony count in the same samples after 24h incubation at 37°C. This indicates that 60 min heat treatment at 70°C and 50% RH effectively inactivates *E. coli* in contaminated N95 respirators.

### Microstructural analysis of the N95 filter layer

To assess whether exposing the polymer microfibers of the N95 filter media to high temperatures caused fiber degradation, we analyzed a 1 cm^2^ filter sample from respirators that underwent 10 thermal disinfection cycles at 70°C and 0% RH or 50% RH. We coated each sample with 10 nm of carbon and imaged with a scanning electron microscope (XL30, FEI company, Oregon, USA) at magnifications of 150x, 200x, 650x, and 1200x at 5 keV. We analyzed fiber morphology with a blinded observer using ImageJ (https://imagej.nih.gov/ij/) in ten randomly selected individual fibers from all quadrants of a representative image of each sample. We measured the fiber diameters in 40 individual fibers per condition (10 fibers per mask type) to calculate the mean fiber diameter after each disinfection cycle.

### Quantitative N95 respirator fit testing

Exposure to high temperatures may affect the mechanical properties of the respirator components, such as elasticity of the headbands or adjustability of the nose clip, potentially allowing leakage of particles. To test the respirator fit, we applied a standardized, quantitative fit testing procedure in a total of 46 respirators (n=12 per type, except n=10 for 1860s) using the a PortaCount Pro+ Respirator Fit Tester 8038 (TSI Incorporated, USA) and a particle generator model 8026 (TSI Incorporated, USA) in compliance with governmental regulatory guidelines (UK Health and Safety Executive, US Occupational Safety and Health Administration [OSHA], and Canadian Standards Association [CSA])(11-13). Particles greater than 0.02 µm in size were detected in a concentration range of 0.01 to 2.5 × 10^5^ particles/cm^3^. Average ambient and in-mask particle concentration was measured during standardized exercises and their ratio was calculated as the respirator fit factor, with a fit factor of 100 being defined by the OSHA as the minimum pass value (11). Fitted respirators were personalized to two blinded test subjects (one male, one female) and underwent thermal disinfection at 0% RH or 50% RH respectively (n=23 each) as outlined above. After 5, 10 and 15 disinfection cycles the quantitative fit testing was repeated for each respirator, with the same test subject. Additionally, the blinded test subject rated the subjective fit, adjustability and comfort of each decontaminated respirator compared to the unprocessed reference masks on the CSA Comfort Assessment Score (0 - no issues; 1 – discomfort can be ignored; 2 – some discomfort but still able to function; 4 – unacceptable discomfort) (13).

### N95 filter efficiency and breathing resistance testing

We determined the breathing resistance and particulate filter efficiency in unprocessed masks (n=12) and in a total of n=58 N95 respirators that underwent either 5 or 10 cycles of thermal disinfection at 0% RH or 50% RH, using the abbreviated National Institute for Occupational Safety and Health (NIOSH) standard (14-16). To measure breathing resistance, the respirators were mounted on a test fixture with air flowing at rate of 85 ±2 l/min. In accordance with the NIOSH standard, a breathing resistance below 343.23 Pa is considered tolerable (16). For NIOSH filtration efficiency protocols, the respirators were pre-conditioned at 85 ± 5% relative humidity and 38 ± 2.5°C for 25 ± 1 hours and then mounted on a certified condensation particle counter (3772, TSI Incorporated, Minnesota, USA). The respirators were tested against a near monodispersed polystyrene latex bead at a flow rate of 85±2 l/min, at 21 - 26°C and 30.4 - 43.2% RH. Particle filter efficiency was calculated as the percentage of all counted particles (median diameter 0.075 ± 0.020 µm) removed by the respirator. For N95 masks, particle filter efficiency needs to be equal to or greater than 95 % (15).

### Statistical methods

We conducted statistical analyses using JMP (version 15.1.0, SAS Institute, USA). Descriptive statistics were calculated among all tested respirators for each condition. All means are expressed with standard deviation (±SD). Given the limited sample size due to the global shortage of N95 respirators, one sample t-tests were used to compare the group means of the disinfected masks to the respective US-regulatory pass value for each assessment. Sample size calculations for quantitative respirator fit revealed a required sample size of n=12 to detect a mean fit factor of 120 (pass value 100, mean fit factor of unprocessed mask 190±15) on an alpha level of 0.01 and a power of 0.95 (17). An alpha level of 0.01 for one-sided *p*-values was chosen to increase the stringency and adjust for multiple comparisons. Additionally, US-regulatory compliance of the disinfected masks was assumed only when the lower bound of the 99% confidence intervals (99% CI) was greater than the minimum required pass value.

## Results

### SARS-CoV-2 inactivation

After inoculation with virus, no infectious SARS-CoV-2 could be detected in any dry heat-treated respirators (70°C for 60 min) whereas high levels of SARS-CoV-2 could still be detected in respirators that did not undergo heat treatment (Table 1).

**Table 1:** SARS-CoV-2 quantification in N95 respirators after thermal disinfection.

| N95 model | Pre-treatment | Titre, Log TCID<sub>50</sub>/ml (mean ± SD) |  |  |
| --- | --- | --- | --- | --- |
|  |  | Control (no dry heat treatment) | 70°C / 0% RH, 60 min* | 70°C / 0% RH, 60min with 5min cool down mid-cycle* |
| 1860S | Unprocessed | 5.62 ± 0.21 | U | U |
|  | 10x heat-treated | 5.69 ± 0.11 | U | U |
| 8110S | Unprocessed | 5.70 ± 0.004 | U | U |
|  | 10x heat-treated | 5.77 ± 0.24 | U | U |
| 8210S | Unprocessed | 5.21 ± 0.50 | U | U |
|  | 10x heat-treated | 5.66 ± 0.08 | U | U |
| 9105S | Unprocessed | 5.56 ± 0.27 | U | U |
|  | 10x heat-treated | 5.45 ± 0.29 | U | U |

\* U: Undetectable; detection limit: 100 Fifty-percent tissue culture infective dose (TCID<sub>50</sub>) per ml
Shown are quantifications of the infective doses of SARS-CoV-2 after a single cycle of thermal disinfection (70°C / 0% RH) of new N95 respirators, and respirators that were pre-treated with 10 disinfections. Infectious SARS-CoV-2 could not be recovered from the disinfected masks, indicating effective decontamination. Increasing the relative humidity during thermal disinfection is therefore not required to inactivate SARS-CoV-2 in N95 respirators.

### Bacterial inactivation

No *E. coli* could be detected in inoculated N95-respirators when heat treated for 60 min at 70°C at 50% RH or at 90°C at 70% RH. As shown in Figure 1, in samples exposed to dry heat (70°C / 0% RH), > 1000 bacterial colonies were still detectable, while exposure to 70°C with humidity (25% RH and 40% RH) dramatically reduced colony formation. Consequently, thermal disinfection for 60 min at 70°C and 50% RH eliminated E. coli contamination on N95 respirators (Table 2).

**Table 2.** **Bacterial quantification in N95 respirators after thermal disinfection using optical density measurement at 600 nm wavelength (OD600) after 24-hour culture**

| Sample No. | Neg Control | Pos Control | 70°C, 0%RH | 70°C, 25%RH | 70°C, 40%RH | 70°C, 50%RH | 90°C, 70%RH |
| --- | --- | --- | --- | --- | --- | --- | --- |
| 1 | 0 | 2.777 | 2.875 | 2.74 | 2.84 | 0 | 0 |
| 2 | 0.001 | 2.851 | 2.861 | 2.625 | 2.768 | 0.04 | 0 |
| 3 | 0.002 | 2.816 | 2.912 | 2.709 | 2.733 | 0.02 | 0 |
| 4 | 0 | 2.64 | 2.736 | 2.748 | 0 | 0 | 0 |
| mean±SD | 0.00±0.00* | 2.77±0.09 | 2.84±0.08 | 2.71±0.06 | 2.09±1.39 | 0.02±0.02* | 0.00±0.00* |

\* significantly different from Pos Control, $p < 0.001$

### Structural properties of the N95 filters

We analyzed the N95 filter media in new, unprocessed (control) respirators and observed an overall mean fiber diameter of 3.88±2 µm. Even after ten cycles of thermal disinfection of 60 min at 70°C and either 0 or 50% RH, the mean overall fiber diameter remained within the range for unprocessed N95 filters as specified in the US patent (Figure 2) (9).

**Figure 2:**
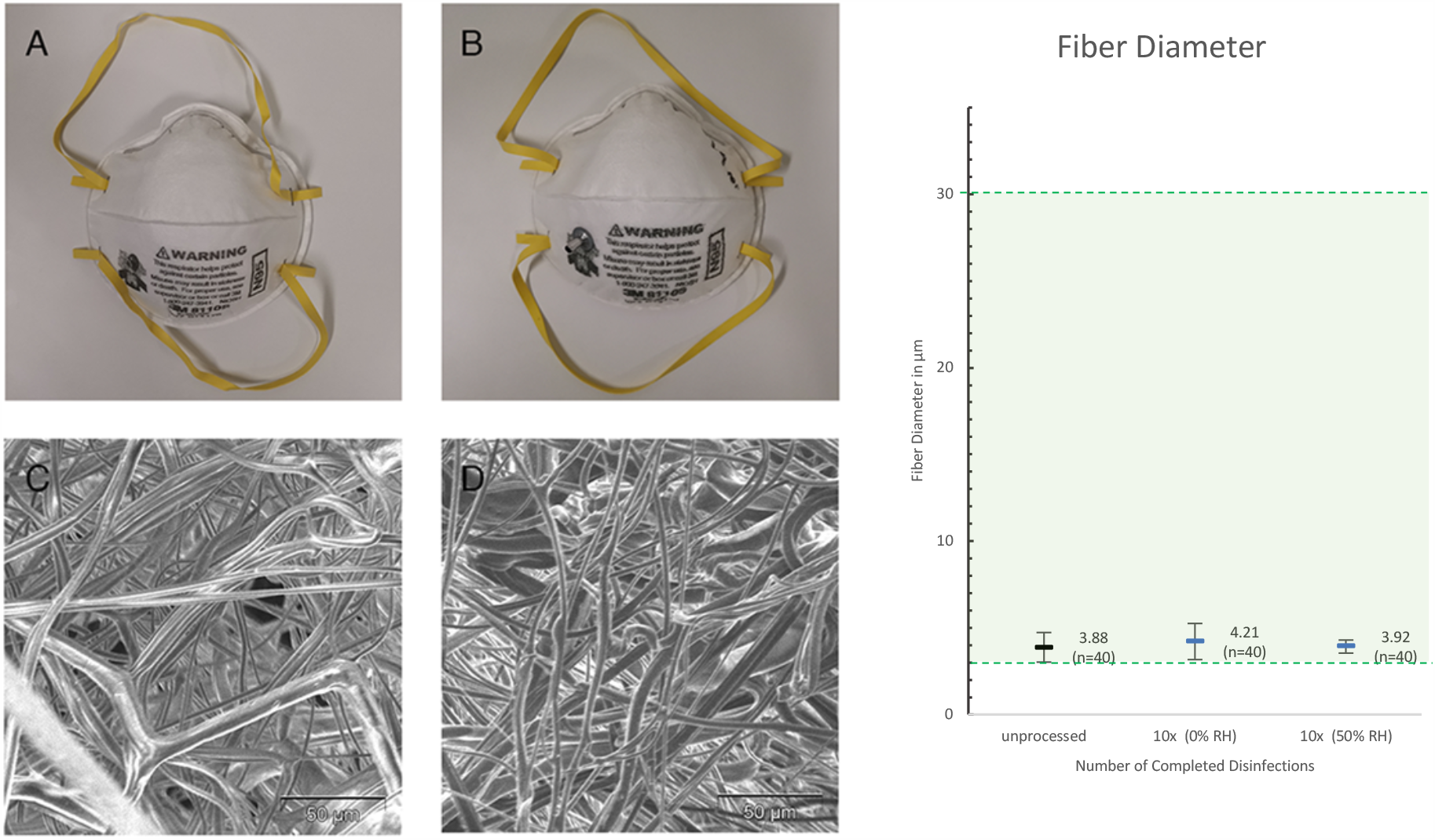
The effect of thermal disinfection on the structural properties of N95 respirators. (Left) The macroscopic and microscopic integrity of the respirator is preserved after repetitive disinfection. The upper left panels compare new (A) to ten times disinfected N95 respirators (B). Panels C and D show their filter layer in 650x magnification (SEM Image, Scale bar 50 µm, type 8110s, 3M, Minnesota, USA). (Right): Shown is the fiber diameter of unprocessed and ten times disinfected N95 filters (0% and 50%RH), with the fiber diameter range of new (untreated) 3M N95 filters shaded in green (9). Shown are the mean fiber diameters with 99% confidence intervals as error bars. Graphs are labelled with mean and sample size.

### Respirator function

Quantitative fit testing was conducted with four common types of commercially available N95 respirators that underwent 5, 10 and then 15 cycles of thermal disinfection at 0% and 50% RH respectively (n=23 for each condition; Figure 3). All tested groups of thermally disinfected respirators significantly exceeded the fit factor of 100, the OSHA-defined standard pass value for sufficient respiratory protection (*p*<0.001 for all groups) and so did the lower bound of their 99% confidence intervals (Figure 3). In a total of 138 performed quantitative fit tests with disinfected respirators (0% and 50% RH), none failed the test. Also, the subjective fit and wearing comfort of the decontaminated respirators did not differ from new masks and were rated with zero, or no issues, on the CSA Comfort Assessment Score.

**Figure 3:**
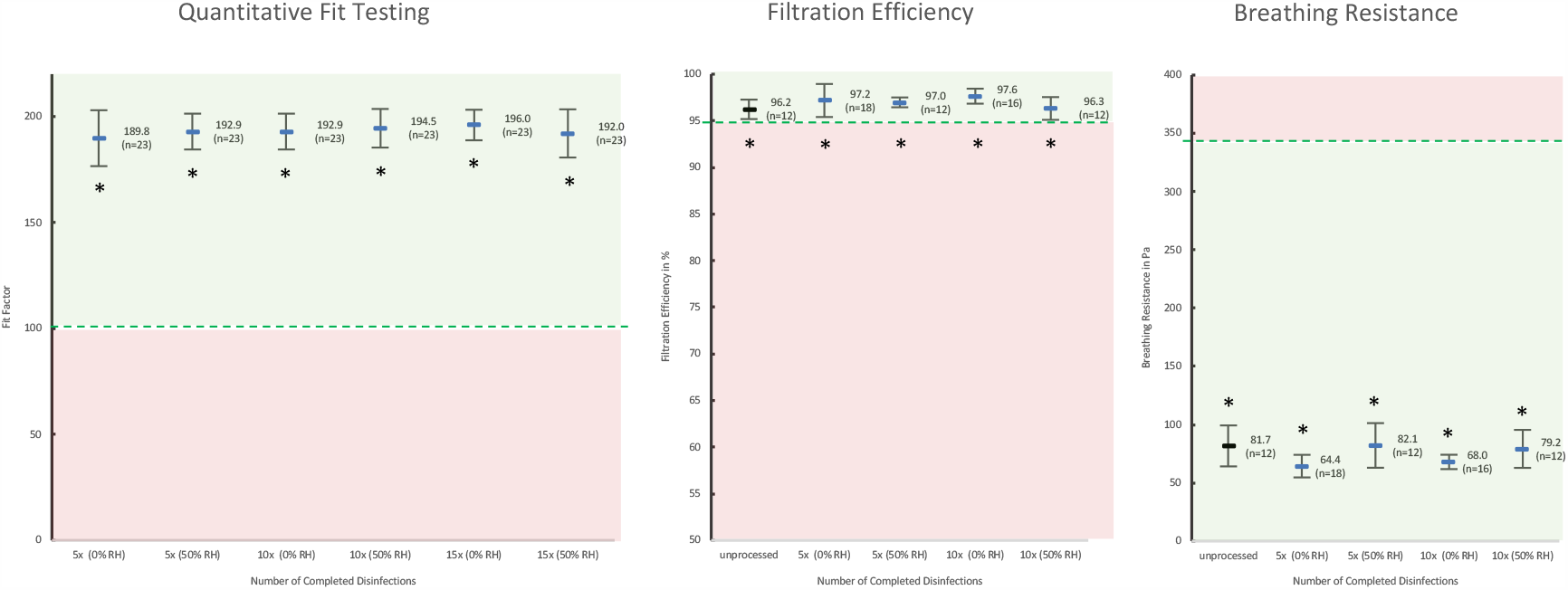
Function of thermally disinfected N95 respirators. Shown are the quantitative fit factors (left), particle filtration efficiency (centre) and breathing resistance (right) of thermally disinfected N95 respirators (0% and 50% RH). The US-governmental pass values for each test are indicated by a dashed line (11, 15, 16). Data are displayed as means with 99% confidence intervals and labelled with the mean and sample size. Groups that significantly exceeded the pass value are labelled with an asterisk (* *p* < 0.01).

Further, we tested the particle filtration efficiency and breathing resistance in the same four types of commercially available N95 respirators that underwent five cycles or ten cycles of thermal disinfection at 0% and 50% RH respectively (Figure 3). The disinfected respirators significantly exceeded 95% filtration efficiency after five and ten disinfection cycles (*p <* 0.001). In addition, the breathing resistance of the same set of disinfected respirators was significantly lower than the maximum tolerable resistance standard of 343.23 Pa for all tested groups (*p* < 0.001).

## Interpretation

Due to an unprecedented demand for personal protective equipment during the COVID-19 pandemic frontline workers are now instructed to re-use disposable N95 respirators (4, 18). Strategies to disinfect and redistribute personalized N95 respirators would increase the safety of health care workers and expand the supply. However, at present, a safe and universally available large-scale decontamination protocol for N95 respirators is unavailable.

Based on recent reports demonstrating the heat sensitivity of SARS-CoV-2, we applied thermal disinfection for four common, disposable N95 respirator models (5, 8). A single disinfection cycle for 60 min at 70°C and 0% RH, effectively inactivated SARS-CoV-2 in all testes respirators. In addition, although airborne bacterial contamination of the N95 respirators to a level representing a respiratory risk for the user may be less concerning in COVID-19 patient care, we also tested for bacterial inactivation: Thermal disinfection thoroughly eliminated *E. coli* in N95 respirators when the relative humidity is kept at 50%, but not below. To further validate the safety of the procedure, we determined the effect of thermal disinfection on the respirator’s integrity and function. Effective filtration of airborne particles relies on both, physical filter pore size of the electret filter layer, and electrostatic attraction (19). The process of disinfection potentially affects these filter properties (20). However, we found that the physical structure of the electret filter media as well as fit, filtration performance and breathing resistance of the N95 respirators is upheld to a level meeting official NIOSH and OSHA standards for at least 10 cycles of thermal disinfection at 0% and 50% RH (11, 15, 16). Also, the subjective wearing comfort is maintained.

In COVID-19 patient care, N95 respirators may be contaminated with virus containing body fluids such as blood, potentially necessitating a longer heat exposure for virus inactivation. To account for that, we increased the exposure time to 60min and don’t recommend reprocessing of visibly contaminated masks. Another potential limitation of the study is that we did not individually test all components of the respirator (e.g. elastic straps) for complete virus inactivation. However, virus within the tested electret filter media is likely to be relatively resistant to heat disinfection compared to other respirator components, suggesting a low risk for elastic straps or other components to remain infective following thermal disinfection.

To effectively cope with the global supply shortage, strategies for disinfection and re-use require widespread scale-up. The U.S. Food and Drug Administration (FDA) recently issued emergency use authorizations for the vaporized hydrogen peroxide gas sterilization of disposable N95 respirators (21, 22). However, this technology is limited to non-cellulose based respirators, therefore making a large proportion of N95s ineligible for reprocessing, and is also unavailable in most hospitals and other facilities (23). Thermal disinfection can be performed at low cost in conventional mechanical convection ovens, which are widely available in commercial kitchens, laboratories or sterilization facilities. Their large capacity enables the simultaneous disinfection of thousands of masks per oven per day, allowing a potential to scale the process to a level sufficient to expand the supply of protective equipment globally. Thereby, thermal disinfection may provide a feasible solution for selected low-and middle-income regions with limited access to personal protective equipment and limited testing capacities, helping to protect their frontline personnel during the COVID-19 pandemic. Other alternative decontamination procedures have been proposed (24-27). Of those, ultraviolet light decontamination systems may represent a promising approach as they seem to maintain respirator function as well (27). However, thorough pathogen elimination, including SARS-CoV-2 in N95 respirators remains to be proven.

In conjunction with such alternative strategies, thermal disinfection may be used as a rapidly applicable emergency measure to alleviate the present global shortage of N95 respirators. Future studies may compare safety, scalability and cost-effectiveness of those decontamination strategies for N95 respirators and specifically investigate SARS-CoV-2 inactivation in respirators contaminated with body fluids such as saliva or blood to further determine safety in real-world conditions.

In conclusion, thermal disinfection for 60 min at 70°C uses widely available equipment to enable the safe reuse of disposable N95 respirators without affecting their protective performance. Given the superior bacterial inactivation, we generally recommend applying 50% humidity during thermal disinfection up to ten times. However, in low-tech/low-income regions it may be challenging to control for humidity. Here, dry heating respirators in mechanical ovens may provide an effective and rapidly scalable low-cost method to thoroughly eliminate SARS-CoV-2 and thereby helps protecting frontline workers from job related risk of infection during the COVID pandemic globally.

## Data Availability

The datasets are available from the corresponding author on reasonable request.

